# Exploring sex-dependent responses to anodal tDCS in subacute stroke rehabilitation: a post-hoc analysis of the randomized sham-controlled NETS Trial

**DOI:** 10.64898/2026.08.19.26360862

**Authors:** Silke Wolf, Linda Krause, Fanny Quandt, Robert Schulz, Anna Suling, Christian Gerloff

## Abstract

**Background:** Upper limb dysfunction is among the most disabling consequences of stroke, yet transcranial direct current stimulation (tDCS), an extensively investigated adjunct to motor rehabilitation, has not demonstrated consistent benefit in large randomized trials. Interindividual variability may contribute, and sex is a plausible source given anatomical and neurophysiological differences relevant to tDCS. In this post-hoc, exploratory secondary analysis of the NETS trial, we examined sex as a potential effect modifier of tDCS response across the 90-day follow-up and all three domains of the International Classification of Functioning, Disability and Health (ICF).

**Methods:** NETS randomized 119 patients with subacute ischemic stroke to anodal tDCS (1 mA) over the ipsilesional primary motor cortex or sham stimulation alongside standardized rehabilitation. Longitudinal mixed-effects models with autoregressive correlation structures examined treatment-by-sex interactions for the Upper-Extremity-Fugl-Meyer Assessment (UEFMA; body function), Box-and-Block Test (BBT; activity), and Stroke Impact Scale participation domain (SIS; participation). Sensitivity analyses included per-protocol analysis, continuous-time models and three-way sex-by-treatment-by-time interactions.

**Results:** Relevant treatment-by-sex interactions were found for the UEFMA and BBT, but not for SIS participation. Women receiving active stimulation improved more than those receiving sham: estimated marginal mean differences: UEFMA 6.0 points (95%-CI 1.2;10.7), BBT 8.4 points (95%-CI 2.4;14.3), whereas no relevant treatment effect was observed in men. Results were similar in intention-to-treat and per-protocol populations.

**Conclusions:** This hypothesis-generating analysis suggests that sex may modify the response to tDCS in post-stroke upper limb rehabilitation. Together with similar subgroup signals from other trials, these findings warrant prospective, adequately powered trials with prespecified tests of treatment-by-sex interaction.

**Highlights:**

- Sex may modify response to transcranial direct current stimulation after stroke
- In this exploratory analysis, women benefited more than men from active stimulation
- Signals of treatment benefit in women were observed for arm function and activity, not for participation
- Sex difference in stimulation response appeared to widen over 90-day follow-up

## 1. Introduction

The promise of transcranial direct current stimulation (tDCS) as an adjunct to post-stroke motor rehabilitation remains unmet, despite more than two decades of clinical investigation [1]. The need for effective adjunct therapies is evident: upper limb dysfunction affects the majority of stroke survivors and is among the strongest determinants of long-term disability [2,3]. Meta-analyses have reported modest positive effects, and there is a plausible rationale for tDCS-induced neuroplasticity. Nevertheless, the large multicenter randomized controlled trials NETS and TRANSPORT2 were neutral on their primary endpoints [4–7]. One likely contributor is substantial interindividual variability in treatment response, which trial designs estimating a single average treatment effect cannot resolve. Biological sex is one candidate source of this variability. High interindividual variability in response is a recognized challenge across the entire field of research on non-invasive brain stimulation. Identifying participant characteristics that explain some of this variability is therefore a priority for this field of research.

Women are markedly underrepresented in clinical stroke research, including studies on non-invasive brain stimulation [8]. Among 34 studies included in a recent Cochrane review on tDCS effectiveness post-stroke, only seven reported a female participation rate of at least 50% [7]. Beyond this underrepresentation, sex differences that may affect tDCS responsiveness are rarely investigated [4,9,10], although there are biological reasons to expect them. Under identical stimulation set-ups, women tend to experience higher current densities than men, a difference further modulated by age [11]. At the same applied current, significantly different electric fields can therefore arise at the cortical target site, and sex could serve as an easily accessible indicator of the anatomical determinants of the delivered dose. Women and men also differ in post-stroke motor cortex physiology: men more often show pronounced interhemispheric asymmetry in motor cortex excitability, whereas women more often maintain balanced interhemispheric activity and may show stronger LTP-like plasticity responses [12–14]. How these differences translate into clinical response to anodal tDCS remains unclear.

However, this question is clinically meaningful, as women experience worse post-stroke outcomes than men [15]. Demographic factors and pre-stroke health account for only approximately 41% of this disparity, leaving a substantial part unexplained [16]. Modifiable factors, such as an individual’s response to neurostimulation, could play a role and therefore require systematic investigation. Both NETS and TRANSPORT2 reported subgroup signals suggesting that tDCS response differs between women and men, although neither trial was adequately powered to detect this difference [4,5]. Moreover, both analyses focused exclusively on impairment-level outcomes, leaving open whether differential effects extend to activity and participation, the domains most relevant to patients’ daily lives. Such reliance on single-domain outcomes, together with the lack of biomarkers to guide patient selection, has recently been highlighted as a key barrier to translation [17]. These questions are particularly relevant in terms of their implications for the planning of future study designs and personalized rehabilitation strategies, especially during the subacute phase, when the potential for neuroplastic interventions may be greatest [10,18].

We therefore conducted a post-hoc, exploratory secondary analysis of NETS, a multicenter, randomized, double-blind, sham-controlled trial of anodal tDCS for upper limb motor recovery in subacute ischemic stroke [5]. Extending the original publication, we examined whether sex modified the effect of tDCS over the full follow-up period and across all three levels of functioning defined by the International Classification of Functioning, Disability and Health (ICF) [19]. These levels were body functions (Upper Extremity Fugl-Meyer Assessment, UEFMA), activity (Box-and-Block Test, BBT) and participation (Stroke Impact Scale, SIS). As this analysis was not prespecified and the trial was not powered to detect treatment-by-sex interactions, it is explicitly hypothesis-generating. Its aim was to determine whether the previously reported subgroup signal is consistent across outcome domains and over time. This would indicate whether sex should be considered as a prespecified effect modifier or stratification factor in future, adequately powered tDCS trials.

## 2. Material and methods

### 2.1 Data and outcomes

The NETS study was conducted at 11 study sites and enrolled patients in the subacute phase of ischemic stroke, defined as 5–45 days after stroke onset. Eligible patients had moderate upper-extremity motor impairment, defined as a UEFMA [20] score of 20–58 points, inclusive, out of a maximum of 66 points, together with either active wrist extension of at least 5° or the ability to perform repetitive grasping movements. Participants were randomized to receive either active anodal tDCS (1 mA, 20 minutes) over the primary motor cortex of the lesioned hemisphere or sham stimulation, administered concurrently with a standardized rehabilitative training program over ten consecutive days. The primary outcome was the change in upper limb motor function, quantified by the UEFMA, measured post-intervention. The detailed methodology, including inclusion/exclusion criteria, randomization procedures, and intervention specifics, has been described comprehensively elsewhere [5,21].

For the current analyses, change from baseline was analyzed at post-intervention (P1, approximately 14 days), and additionally at follow-up assessments at 30 (FU1) and 90 (FU2) days after randomization. In addition to the primary outcome of the initial analysis (change in UEFMA at P1), secondary endpoints included the change in the BBT [22], which assesses manual dexterity at the activity level, and in the participation domain of the SIS [23]. These additional endpoints enable a comprehensive evaluation of functioning across the three levels of the ICF framework. Missing values were handled using the Last-Observation-Carried-Forward approach.

All analyses were performed within the intention-to-treat (ITT) population, defined as all randomized subjects who received at least one stimulation session, and subsequently within the per-protocol (PP) population, which excluded subjects with major protocol deviations. All patients were analyzed according to their randomized treatment assignment.

### 2.2 Statistical analysis

The procedure and steps of the analysis are illustrated in Figure 1. Baseline characteristics are presented across the four subgroups defined by sex and treatment allocation (women sham, women verum, men sham, men verum). Continuous variables were summarized as median and interquartile range, categorical variables were reported as frequencies and percentages.

**Figure 1:**
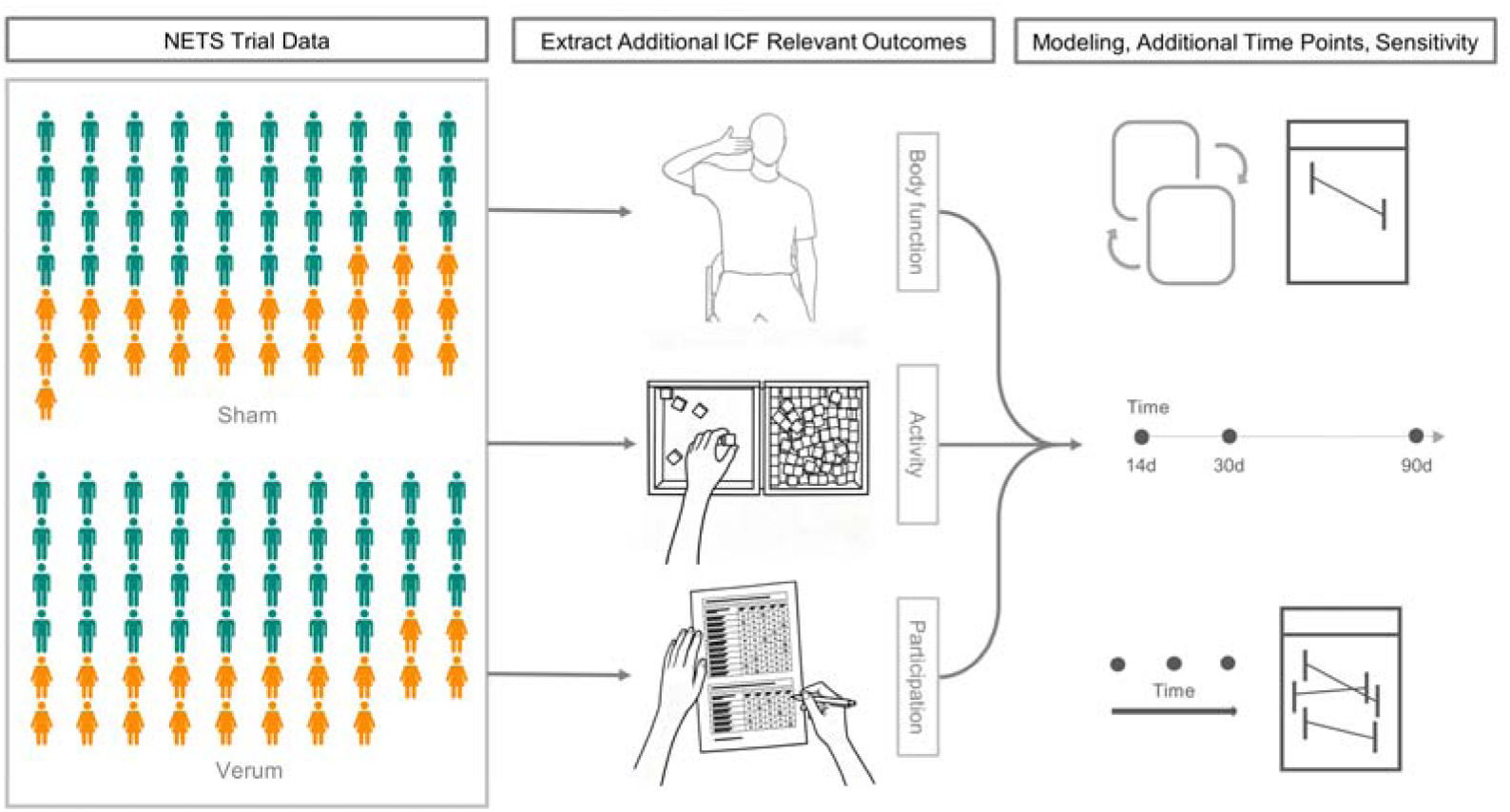
Procedure for the Exploratory Post-Hoc Analysis

For the original NETS analysis, the effect of treatment was estimated using a linear model. This model estimated the effect of treatment (verum vs. sham) on the change in UEFMA from baseline to post-intervention (P1):

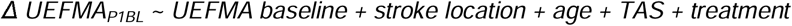

where the fixed effects included the baseline score, time after stroke (TAS) in days, and treatment group, as well as randomization stratification factors: stroke location (cortical vs. sub-cortical) and age. Subsequently, the original subgroup analysis included sex and its interaction with treatment:

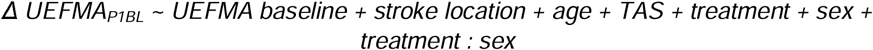

Building on these basic models, the current analysis fitted extended mixed linear models, which additionally account for longitudinal changes, for each of the three ICF outcome measures, using the nlme package [24]. These models incorporated repeated measurements across all follow-up time points after randomization (post-intervention: P1, approximately 14 days, and additionally at follow-up assessments at 30 days: FU1, and 90 days: FU2) to assess how effects evolve over the recovery period. Residual correlations over time within individuals were modeled using an autoregressive AR(1) structure. The respective baseline values, stroke location, age, and TAS were included as covariates (the term FU refers collectively to all follow-up assessments: P1, FU1 and FU2):

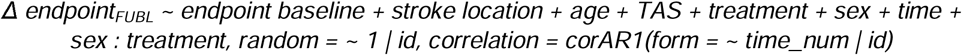

Interactions were tested using Type II Wald chi-square tests via the car package [25]. Estimated Marginal Means (EMMs) were derived using the emmeans package [26] to facilitate interpretation of treatment-by-sex differences, with pairwise comparisons used to assess specific contrasts within each subgroup.

Sex was examined as a potential effect modifier by testing the interaction between treatment and sex, rather than comparing treatment effects separately within each sex. Testing separately within the subgroups would not provide information on whether the treatment effect differs between the subgroups, even if both were statistically significant [27]. The treatment effects in women and men are presented as effect estimates to illustrate the direction and magnitude of the interaction.

Additional sensitivity analyses were performed. First, models with time as a continuous predictor were fitted to evaluate the linearity assumption of recovery trajectories. Second, higher-order interactions (sex × treatment × time) were incorporated to explore potential differences in response patterns between men and women across the follow-up period:

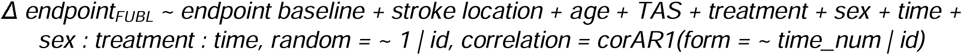

Third, the models were further adjusted for the overall severity of the stroke at the start of the study to assess the robustness of the effect estimates, with the NIHSS score at baseline included as an additional covariate.

Residuals were examined through homoscedasticity plots, Q-Q plots, and histogram distributions to verify assumptions of normality and constant variance. All analyses were conducted in the software R, version 4.4.1 [28].

## 3. Results

The baseline demographic and clinical characteristics of the study participants are summarized in Table 1, with detailed information on the distribution of risk factors provided in the supplementary material (Table S1). The total sample comprised 119 participants in the ITT population, including 24 women and 37 men in the sham group, as well as 20 women and 38 men in the verum group. The PP population consisted of 94 participants, with 21 women and 30 men in the sham group, and 17 women and 26 men in the verum group.

**Table 1:** Baseline Demographic and Clinical Data.

|  | **Sham Women (N=24)** | **Sham Men (N=37)** | **Verum Women (N=20)** | **Verum Men (N=38)** |
|---|---|---|---|---|
| **Age (y)** | 74 (66-80) | 65 (57-71) | 68 (56-76) | 66 (60-73) |
| **TAS (d)** | 22 (10-28) | 18 (9-26) | 20 (12-36) | 21 (10-25) |
| **Localization** |  |  |  |  |
| **Cortical** | 9 (38) | 13 (35) | 5 (25) | 14 (37) |
| **Subcortical** | 15 (62) | 24 (65) | 15 (75) | 24 (63) |
| **NIHSS** | 4 (3-5) | 3 (2-4) | 4 (3-5) | 4 (3-5) |
| **MMSE** | 29 (26-30) | 29 (28-30) | 29 (27-30) | 29 (26-30) |
| **BBT** | 16 (2-30) | 20 (6-28) | 15 (0-20) | 12 (2-24) |
| **SIS Participation** | 45 (25-70) | 50 (22-69) | 34 (25-48) | 25 (11-49) |
| **SIS Recovery** | 32 (14-50) | 35 (30-50) | 45 (40-50) | 30 (22-50) |
| **UEFMA** | 38 (26-48) | 42 (32-50) | 34 (29-50) | 36 (28-44) |

BBT – Box-and-Block Test, d – days, MMST – Mini Mental State Examination, NIHSS – National Institutes of Health Stroke Scale, SIS – Stroke Impact Scale, TAS – Time After Stroke, UEFMA – Upper-Extremity-Fugl-Meyer Assessment, y – years.
Results reported as Median (IQR) \| n (%)

The results of the original analysis are presented in the supplement. Following the analytical framework of the original NETS publication, no evidence of an overall treatment effect was found in the total sample (change verum vs sham −0.3, 95%-CI [−3.0; 2.4]; *p=0.820*, Table S2). The original exploratory subgroup analysis suggested greater improvements in UEFMA scores among women receiving verum compared to those receiving sham (mean treatment difference in women: 4.6, 95%-CI: 0.2; 9.0). No relevant differences were observed between treatment groups for men (Table S3).

Based on these results from the original analyses, the expanded linear mixed-effects models examined sex-specific effects and recovery trajectories over the full follow time for all outcomes. The results indicated treatment-by-sex interactions for the UEFMA and the BBT (the estimated between-group differences are shown in Table 2). Women in the verum group showed greater improvements compared to the sham group, with an estimated difference of 6.0 points (95%-CI: 1.2;10.7) for the UEFMA, and 8.4 points (95%-CI: 2.4;14.4) BBT. In contrast, no notable differences between treatment groups were observed in men for these measures, with estimated differences of -2.2 points (95%-CI: -5.7;1.3) for the UEFMA, and - 3.4 points (95%-CI: -7.9;1.1) for the BBT. For the SIS participation, no relevant treatment-by-sex interaction was observed (Table 2, Figure 2).

**Figure 2:**
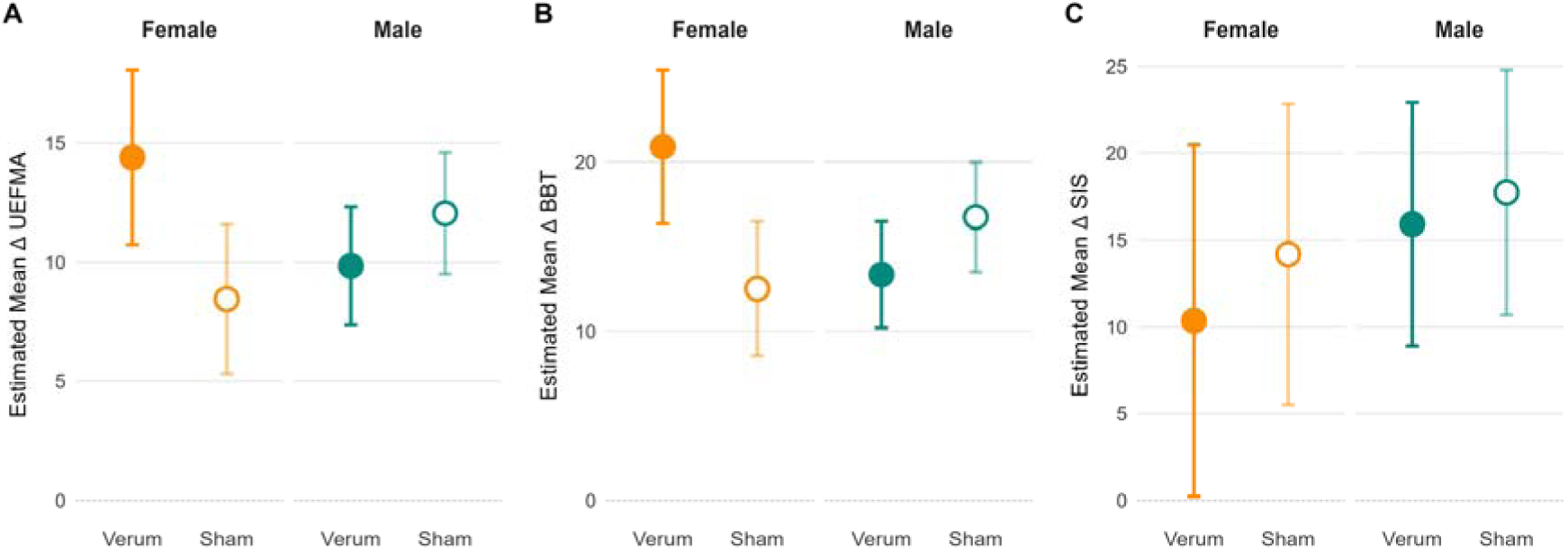
Estimated Marginal Means (95%-CI) for Change Scores by Treatment Group and Sex (values for women displayed in orange, for men in teal). A – Upper-Extremity-Fugl-Meyer Assessment (UEFMA), B – Box-and-Block Test (BBT), C – participation domain of the Stroke Impact Scale (SIS).

**Table 2:** Results from Expanded Models on Additional Endpoints and Increased Follow-up Time.

| **Model** | **Regression Coefficient** | **95%-CI** |  | ***p*** |
|---|---|---|---|---|
| **UEFMA** |  |  |  |  |
| Interaction treatment : sex |  |  |  | 0.022 |
| Verum vs. Sham (women) | 6.0 | 1.2 | 10.7 | 0.015 |
| Verum vs. Sham (men) | -2.2 | -5.7 | 1.3 | 0.218 |
| Baseline UEFMA | -0.4 | -0.5 | -0.3 | <0.001 |
| Location (subcortical vs. cortical) | 0.2 | -2.8 | 3.1 | 0.905 |
| Age | -0.2 | -0.3 | -0.1 | 0.002 |
| TAS | -0.3 | -0.4 | -0.2 | <0.001 |
| Time (FU1 vs. P1) | 2.7 | 1.5 | 3.8 | <0.001 |
| Time (FU2 vs. P1) | 3.5 | 2.3 | 4.6 | <0.001 |
| **BBT** |  |  |  |  |
| Interaction treatment : sex |  |  |  | 0.007 |
| Verum vs. Sham (women) | 8.4 | 2.4 | 14.4 | 0.006 |
| Verum vs. Sham (men) | -3.4 | -7.9 | 1.1 | 0.135 |
| Baseline BBT | -0.2 | -0.3 | -0.1 | 0.003 |
| Location (subcortical vs. cortical) | 3.2 | -0.6 | 6.9 | 0.094 |
| Age | -0.3 | -0.5 | -0.2 | <0.001 |
| TAS | -0.4 | -0.5 | -0.2 | <0.001 |
| Time (FU1 vs. P1) | 6.2 | 4.7 | 7.7 | <0.001 |
| Time (FU2 vs. P1) | 10.0 | 8.5 | 11.6 | <0.001 |
| **SIS participation** |  |  |  |  |
| Interaction treatment : sex |  |  |  | 0.798 |
| Verum vs. Sham (women) | -3.8 | -17.2 | 9.5 | 0.569 |
| Verum vs. Sham (men) | -1.8 | -11.8 | 8.1 | 0.716 |
| Baseline SIS participation | -0.6 | -0.7 | -0.4 | <0.001 |
| Location (subcortical vs. cortical) | -5.1 | -13.2 | 3.0 | 0.216 |
| Age | -0.2 | -0.5 | 0.2 | 0.348 |
| TAS | -0.3 | -0.6 | 0.0 | 0.083 |
| Time (FU1 vs. P1) | 9.7 | 5.4 | 14.0 | <0.001 |
| Time (FU2 vs. P1) | 18.3 | 14.0 | 22.7 | <0.001 |

BBT – Box-and-Block Test, SIS – Stroke Impact Scale, TAS – Time After Stroke, UEFMA – Upper-Extremity-Fugl-Meyer Assessment.

In the analyses of the PP population, the same treatment-by-sex interactions were found to be relevant for UEFMA and BBT, with numerically larger effect estimates (Table S4).

The modeling of the factor time as a continuous variable instead of a categorical variable had no relevant impact on the effect estimates, except for the specific estimates regarding time (Table S5). Nonetheless, time remained an independent predictor of recovery across all three outcomes (Figure S1).

Further sensitivity analyses, including the three-way interaction analysis between treatment, sex, and time, were performed. No distinct patterns of interaction were observed (Table S6). However, there was a tendency that the functional differences between men and women become more pronounced over time, most notably for the BBT (Figure 3). Additional adjustment for baseline NIHSS did not substantially change the treatment effect estimates. For the UEFMA, the treatment effect in women was 5.9 points (95%-CI 1.1; 10.7) and in men −2.3 (95%-CI −5.8; 1.3). Likewise, the effects for the BBT (women: 8.4, 95%-CI 2.4; 14.4; men: −3.4, 95%-CI −7.9; 1.1) and the SIS (women: -3.6, 95%-CI -16.9; 9.7; men: −1.1, 95%-CI −11.1; 8.9) remained nearly unchanged.

**Figure 3:**
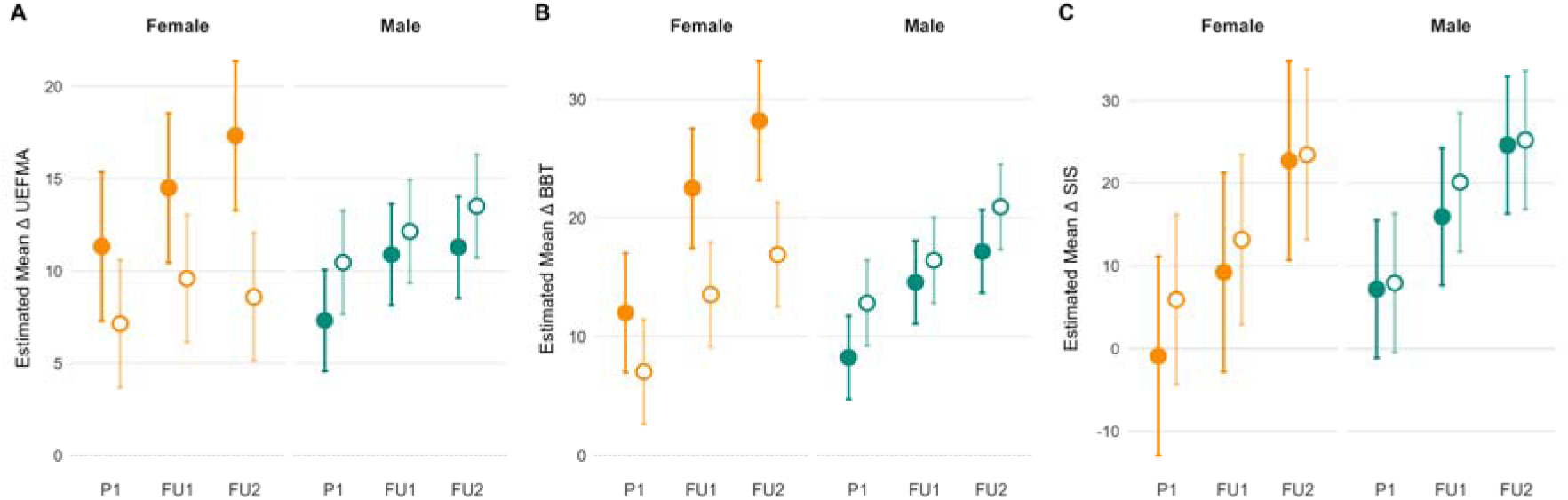
Estimated Marginal Means (95%-CI) for Change Scores from Triple-Interaction by Treatment Group, Sex, and Time (values for women displayed in orange, for men in teal). A – Upper-Extremity-Fugl-Meyer Assessment (UEFMA), B – Box-and-Block Test (BBT), C – participation domain of the Stroke Impact Scale (SIS).

## 4. Discussion

### 4.1 Summary of main findings

This post-hoc exploratory secondary analysis of the NETS trial outcomes examined sex as a potential effect modifier of tDCS response in upper limb rehabilitation during the subacute phase post-stroke. The analyses suggested a potential treatment-by-sex interaction in two ICF domains: female participants showed larger improvements with active tDCS than with sham for both body function (6.0-point UEFMA difference) and activity (8.4-point BBT difference), whereas no relevant treatment effect was observed in male participants for either measure. No relevant differential effect was found for the SIS participation domain. The pattern was similar in both the ITT and PP populations. In the active group, the difference between women and men appeared to increase over the follow-up period, particularly for the BBT. Given the post-hoc design and the small subgroups, these findings should be regarded as hypothesis-generating rather than as evidence of a sex-specific treatment effect.

### 4.2 Clinical relevance and convergence

The observed potential sex-specific effects are consistent with subgroup signals from other trials. In TRANSPORT2, estimated mean UEFMA improvements were numerically larger in female than male participants [4] . Balikshahi et al. reported that neurological, but not motor-specific, improvement with bihemispheric tDCS was greater in female patients and those under 60 years of age [29]. That a similar directional signal emerges across trials differing in stimulation intensity, montage, stroke phase, and concurrent therapy is compatible with an effect that is not protocol-specific. The estimated marginal mean differences of approximately 6 points for the UEFMA and 8 points for the BBT in the current analyses approach thresholds of clinically important differences in moderately affected patients [30,31]. If confirmed, effects of this magnitude would be clinically relevant.

The pattern across ICF domains is also of interest: a potential interaction was observed for the UEFMA and BBT but not for SIS participation. This is consistent with the broader tDCS literature showing diminishing evidence across ICF levels [7,32]. This limitation has recently been reiterated as a barrier to translation [17]. Several items in the participation assessments depend only to a limited extent on bilateral upper extremity function; moreover, this area of function depends on multiple other variables. Thus, participation is shaped by non-motor determinants that tDCS does not directly address. This dissociation is particularly relevant given that women experience persistently worse participation-level outcomes after stroke – a disparity that cannot be assessed with impairment-level measures alone and that motor interventions alone are unlikely to resolve, underscoring the need for participation-specific outcomes and complementary interventions in future trials [15,33,34].

### 4.3 Mechanistic considerations

Several mechanisms could plausibly contribute to the observed pattern, although none could be tested directly in this analysis. Anatomically, sex differences in cranial bone composition and brain torque result in differential current delivery under identical stimulation parameters [35,36]. Modeling studies indicate that women receive higher current densities at the cortex than men. This is in part because greater scalp-to-cortex distance and skull thickness in men attenuate the electric field reaching the target region, meaning that a fixed stimulation intensity may be effectively subtherapeutic in male patients. From a neurophysiological perspective, men tend to exhibit greater interhemispheric asymmetry after a stroke, while women are more likely to maintain balanced interhemispheric activity and demonstrate longer-lasting neuroplastic responses [12–14,37]. Estrogen-mediated enhancement of synaptic plasticity has been described in younger women, but its relevance to the predominantly postmenopausal population studied here is uncertain [38].

The temporal pattern of the observed effects is particularly noteworthy. The functional difference between women and men in the active group appeared to become more pronounced over the 90-day follow-up, most notably for the BBT, although no clear three-way interaction pattern emerged. If genuine, this widening gap would suggest that the potential sex-specific effect reflects a sustained divergence in neuroplastic response rather than a transient difference in early recovery. This interpretation would be consistent with the notion that women may show longer-lasting neuroplastic responses to stimulation. Such a pattern would carry implications for the optimal duration of follow-up in future trials, as effects that emerge gradually may be missed by assessments limited to the immediate post-intervention period. Given the exploratory nature of this analysis, however, this observation remains strictly hypothesis-generating.

Taken together, these potential mechanistic pathways (anatomical, neurophysiological, and potentially hormonal) provide a plausible biological framework for the hypothesis of a sex-dependent treatment response.

### 4.4 Limitations and future directions

Several limitations should be taken into account when interpreting the present results. This is a retrospective, exploratory secondary analysis, and the interaction between treatment and sex was not a predefined primary hypothesis, which entails an increased risk of a Type I error. The subgroups are small, particularly among women in the active treatment group (n = 20 ITT, n = 17 PP), which limits statistical precision and generalizability. For the same reason, the absence of an effect among men or in the area of participation cannot be interpreted as evidence of a lack of effect. Accordingly, these results require prospective confirmation in a study with sufficient statistical power. No hormonal data were collected, precluding direct assessment of estrogen’s contribution. Individual anatomical data were not available for electric field modelling, preventing disentanglement of anatomical from neurobiological sources of sex differences, a gap that individualized computational modelling in prospective studies may help to address [39].

The NETS trial used a single stimulation intensity of 1 mA, and whether the sex-specific effects generalize to higher intensities or alternative montages remains unknown. The Last Observation Carried Forward approach, while consistent with the original pre specified NETS analysis, may introduce bias if dropout patterns differ between subgroups. Finally, key variables including corticospinal tract integrity, baseline cognitive function, and neglect, all established predictors of tDCS response, were not available for analysis, and their potential interaction with sex remains unexplored [6].

If confirmed, the difference in effect size between the sexes would have implications for future trial design. Trials estimating only an average treatment effect across a heterogeneous population may fail to detect an effect that is confined to one subgroup. Confirming a sex-dependent effect would require prespecified tests of the treatment-by-sex interaction in trials adequately powered for this purpose, ideally with sex-stratified randomization to ensure balanced allocation. Separate within-sex comparisons are not sufficient, as they do not test whether the treatment effect differs between sexes [27]. Understanding the mechanistic basis of such differences would additionally require the collection of biological variables such as hormonal status, anatomical data for electric field modelling, and corticospinal tract integrity. These priorities align with the SRRR3 translational roadmap, which called for greater individualization of NIBS protocols and standardization of ICF-aligned outcome measurement [40].

## 5. Conclusions

This exploratory analysis suggests a potential treatment-by-sex interaction regarding response to tDCS in post-stroke upper limb rehabilitation. The potential treatment-by-sex interaction extended across two ICF domains and was consistent across analytical populations, with effect estimates in women approaching clinically important thresholds. Together with results from other studies, these findings suggest that the neutral primary endpoints of large tDCS trials may partly reflect the dilution of a sex-specific treatment effect in unstratified study populations. Prospectively powered trials with sex-stratified randomization and analysis, biological and anatomical assessments, and outcomes spanning all ICF domains are needed to test this hypothesis and advance toward personalized brain stimulation protocols for stroke rehabilitation.

## Non-standard Abbreviations and Acronyms

BBT: Box-and-Block Test
ITT: Intention-to-treat population
ICF: International Classification of Functioning, Disability, and Health
MMST: Mini Mental State Examination
NIHSS: National Institutes of Health Stroke Scale
PP: Per-protocol population
SIS: Stroke Impact Scale
TAS: Time after stroke
tDCS: Transcranial direct current stimulation
UEFMA: Upper-Extremity-Fugl-Meyer Assessment

## CRediT authorship contribution statement

**Silke Wolf**: Writing – original draft, Writing – review & editing, Conceptualization, Data curation, Formal analysis, Funding acquisition, Methodology, Project administration, Visualization. **Linda Krause**: Writing – review & editing, Data curation, Methodology, Validation. **Fanny Quandt**: Writing – review & editing, Investigation, Resources, Supervision. **Robert Schulz**: Writing – review & editing, Investigation, Resources, Supervision. **Anna Suling**: Writing – review & editing, Data curation, Methodology, Validation. **Christian Gerloff**: Writing – review & editing, Funding acquisition, Resources, Supervision.

## Ethics declaration

The trial protocol was approved by the relevant national or local ethics committees or institutional review boards at all participating sites. Written informed consent was obtained from all the patients.

## Data availability statement

The anonymized data that support the findings of this study are available from the corresponding author upon reasonable request.

## Declaration of generative AI

During the preparation of this work the authors used Claude (Claude Opus 5.5, Anthropic) for editing language and stylistic aspects, and to generate suggestions for phrasing and code, including code review. The tool was not used for study design, data analysis, or the interpretation of results. After using this tool, the authors reviewed and edited the content as needed and take full responsibility for the content of the published article.

